# The General Psychopathology Factor from Early to Middle Childhood: Longitudinal Genetic and Risk Analyses

**DOI:** 10.1101/2021.03.20.21253838

**Authors:** Reut Avinun, Ariel Knafo-Noam, Salomon Israel

## Abstract

Accumulating research suggests the structure of psychopathology is best represented by continuous higher-order dimensions, including a general dimension, *p*-factor, and more specific dimensions, e.g., residualized externalizing and internalizing factors. Here, we aimed to 1) replicate *p* in early childhood; 2) externally validate the factors with key constructs of psychological functioning; 3) examine stability and change of genetic and environmental influences on the psychopathology factors from early-to mid-childhood; 4) examine the factors’ predictive utility; and 5) test whether the factors can be predicted by early life measures (e.g., neonatal complications). The Longitudinal Israeli Study of Twins from age 3 to 9 was used for the analyses. Mothers reported on developmental problems, pregnancy and neonatal conditions, and filled in questionnaires on each twin’s externalizing and internalizing symptoms. Cognitive ability was assessed in the lab at age 6.5 and personality traits, self-esteem, and life satisfaction were self-reported by the twins at ages 11-13. A bifactor model that included *p* and externalizing and internalizing factors fit the data best and associations between *p*, cognitive ability, and personality were replicated. Longitudinal twin analyses indicated that *p* is highly heritable (64-73%) with a substantial proportion of the genetic influences stable from age 3. The residualized internalizing and externalizing factors were also highly heritable. Higher *p* predicted developmental problems at age 8-9 and lower self-esteem at age 11. Early life measures were not strongly associated with psychopathology. Our results show that *p* is discernible in early childhood, highly heritable, and prospectively associated with negative outcomes.

**General Scientific Summary:** The general psychopathology factor is discernible in early childhood, highly heritable, with genetic influences contributing to both stability and change, and prospectively associated with developmental problems and lower self-esteem. Early life measures, such as birth complications or hospitalizations during the first year of life, were not strong predictors of the general psychopathology factor or the residualized externalizing and internalizing factors.

---

Comorbidity of psychopathology is prevalent. About 40% of individuals with one class of disorders (e.g., mood, anxiety, substance abuse) are likely to be diagnosed with another (Merikangas et al., 2010; Newman, Moffitt, Caspi, & Silva, 1998). As a result of this co-occurrence, instead of discrete mental disorders, researchers have begun to conceptualize the structure of psychopathology as consisting of broad transdiagnostic dimensions, including externalizing (e.g., hyperactivity and antisocial behavior), internalizing (e.g., depression and anxiety), and thought disorders (schizophrenia and obsessive compulsive disorder). Accumulating evidence has suggested that in addition to specific factors, such as externalizing and internalizing, a general psychopathology factor, often called *p* (Caspi et al., 2014), is also needed to represent the structure of psychopathology (Lahey et al., 2012). The *p*-factor captures shared variation across most, if not all, types of psychopathology and is thought to represent a general liability for mental disorders, as well as their persistence and severity (Caspi & Moffitt, 2018).

The *p*-factor has been identified in various age groups, from preschoolers to adolescents and adults; in clinical and general population samples; in different countries; and using different methods and measures (e.g., Caspi et al., 2014; Gomez, Stavropoulos, Vance, & Griffiths, 2019; Hyland et al., 2018; Laceulle, Chung, Vollebergh, & Ormel, 2020; Olino, Dougherty, Bufferd, Carlson, & Klein, 2014; Patalay et al., 2015; Snyder, Young, & Hankin, 2017). To characterize and validate the *p*-factor, researchers have examined its association with broadly informative indicators of psychological functioning such as personality traits and cognitive ability. In early and middle adulthood, the *p*-factor has been found to correlate positively with neuroticism and negatively with agreeableness and conscientiousness (e.g., Avinun, Romer, & Israel, 2020; Caspi et al., 2014; Etkin, Mezquita, López-Fernández, Ortet, & Ibáñez, 2020). Correlations between the *p*-factor and measures relating to cognitive ability and executive function have been found to be negative in middle childhood (Martel et al., 2017), early-adolescence (Patalay et al., 2015), and adulthood (Caspi et al., 2014).

Further supporting the *p*-factor’s reliability are genetic studies showing that the *p*-factor is genetically influenced, i.e., not only explained by noise and measurement error, and that these genetic influences are relatively stable across time (Allegrini et al., 2020; Riglin et al., 2019; Selzam, Coleman, Caspi, Moffitt, & Plomin, 2018). A longitudinal twin analysis from childhood (age 7 years) to adolescence (age 16 years), has shown that the *p*-factor is substantially heritable (50-60%) across ages and that the genetic component is relatively stable, so that the genetic influence at age 7 accounts for most of the genetic variance in later ages (Allegrini et al., 2020). Notably, to our knowledge, the latter study is the only study to date that employed a longitudinal twin design to examine change and stability in the genetic and environmental effects on the *p*-factor, and no study has examined the specific/residualized factors (e.g., externalizing and internalizing). Additional developmental research is therefore warranted, especially during early life when executive functions, which are significantly linked to externalizing and internalizing symptoms (Hughes & Ensor, 2011), rapidly develop.

The *p*-factor has also been shown to predict negative outcomes (e.g., Blanco et al., 2019; Lahey et al., 2015). For example, estimates of the *p*-factor in middle childhood and early adolescence predict psychiatric diagnoses (anxiety, depression, and alcohol and drug use disorders), use of anxiolytics and antidepressants, school failure, and court convictions in adolescence (Pettersson, Lahey, Larsson, & Lichtenstein, 2018). Taken together, the above findings support the reliability and validity of the *p*-factor as a common variance that captures an inherent and general risk for psychopathology.

Although the shared variance across mental disorders is a robust and replicable finding, the mechanisms that underlie this general risk are less clear and longitudinal studies are scarce. While various candidates likely play a role in the etiology of the *p*-factor, of special interest perhaps, are those that are present at or around birth, as previous studies suggest that the *p*-factor can be found early in development, in children as young as 4 years old (Morales et al., 2021). For example, the link between the *p*-factor in adulthood and childhood socioeconomic status (SES) has been examined, as studies have shown that children raised in families with low SES show increased risk for psychopathology (Peverill et al., 2020), particularly externalizing behaviors. Interestingly, at least one study has found the correlation between the *p*-factor and childhood SES to be weak (Caspi et al., 2014).

Pregnancy, obstetric, and neonatal complications have previously been identified as increasing risk for poorer mental health and/or poorer cognitive function (De Mola, De França, de Avila Quevedo, & Horta, 2014; Ehrenstein et al., 2009; Hamlyn, Duhig, McGrath, & Scott, 2013; Van Lieshout & Voruganti, 2008). For example, Chiorean and colleagues (2020) found that children admitted to neonatal/special care units at birth were more likely to develop a psychiatric disorder in adolescence. Similarly, preterm birth has been associated with increased risk of psychiatric hospitalizations in young adulthood (Nosarti et al., 2012), and pregnancy/birth complications (e.g., prenatal bleeding, hypertension and diabetes, breech birth, and instrument delivery) have been associated with externalizing problems at age 11 (Liu, Raine, Wuerker, Venables, & Mednick, 2009). Because these factors are not necessarily specific to any particular psychopathology, they may serve as transdiagnostic risk factors.

In the current study, we test the replicability of the general psychopathology factor finding from early to middle childhood (e.g., Hankin et al., 2017; Olino et al., 2014), on a longitudinal sample of twins followed from age 3 to age 9. Next, we evaluate the external validity of *p* and the specific/residualized externalizing and internalizing factors by testing for associations with personality traits and cognitive ability, and examine if/how these associations change during development. Following these analyses, we conduct a genetically informed longitudinal analysis from early to middle childhood to examine stability and change of the genetic and environmental influences on the psychopathology factors. This analysis can provide further support for the reliability of the factors by showing their stability across time. We next tested the predictive value of the three factors in the context of developmental problems at age 8-9 (whether the child suffers from e.g., stammering, speech delay, and emotional, social, or communication problems), self-esteem at age 11, and life satisfaction at age 13. Finally, we conducted exploratory analyses to examine whether measures that can be assessed early in the child’s life, such as SES at age 3 and pregnancy, obstetric, and neonatal complications, can predict the psychopathology factors. We hypothesized that risk measures, such as low SES, low birth weight and complications of pregnancy (e.g., rubella, RH mismatch, bleeding during pregnancy or miscarriage risk, and preeclampsia), will be associated with higher scores of the psychopathology factors.

## Methods

### Participants

Families in this study were participants in the Longitudinal Israeli Study of Twins (LIST), a study of social development, in which parents of all Hebrew-speaking families of twins born in Israel during 2004-2005 were invited to participate (Avinun & Knafo, 2013; Vertsberger, Abramson, & Knafo-Noam, 2019). At each wave the experimental protocol was approved by the ethics committee of either the university or the local hospital, and informed consent was obtained from all participating parents. Mothers were asked to complete questionnaires regarding their pregnancy (only age 3 and 5), demographic details, and children’s development, when the twins were 3, 5, 6.5, and 8-9 years old. Data for this study was available for 2,770 children at age 3 (M=3.16 years, SD=.26; 50.3% males; 582 MZ [monozygotic], 2,164 DZ [dizygotic], and 24 of unknown zygosity), 1,908 at age 5 (M=5.13 years, SD=.14; 51% males; 370 MZ, 1,518 DZ, and 20 of unknown zygosity), 1,020 at age 6.5 (M=6.57 years, SD=.30; 50% males; 285 MZ, 729 DZ, and 6 of unknown zygosity), and 973 at age 8-9 (M=9.01 years, SD=.52; 48.7% males; 260 MZ, 699 DZ, and 14 of unknown zygosity). Due to design and budgetary constraints, at ages 6.5 and 8-9 only a small number of opposite-sex DZ twins and only families with which it was possible to schedule an in-person experiment were recruited. Twins’ zygosity was determined based on either a DNA analysis or a parental questionnaire of physical similarity, which has been shown to be in 95% agreement with DNA information (Goldsmith, 1991).

About 25% of the age 3 families did not participate in later waves, a comparison of this group of families to families that did participate in at least one more wave is shown in Supporting Table 1. There were no significant differences in the means of SES or mother reports of children’s psychopathology (the assessment of these measures is detailed below. Comparisons were made in Mplus version 7, using the type=complex with the ‘cluster’ option, to account for the dependent structure of the data).

### Measures

#### Psychopathology

Similar to a previous study in childhood that relied on symptom-level, instead of disorder-level, scores (Patalay et al., 2015), items from 2 different questionnaires were used for the assessment of psychopathology. Internalizing symptoms were assessed in the four waves by mothers’ ratings of 5 items from the emotional symptoms subscale of the Strengths and Difficulties Questionnaire (SDQ; Goodman, 1997) and 5 items from the negative emotionality subscale of the Emotionality, Activity and Sociability Temperament Survey (Buss & Plomin, 1984). Ratings were given on a 3-point scale ranging from 0 (not true/rare) to 2 (very true/often) or on a 5-point scale ranging from 1 (does not characterize at all) to 5 (highly characterizes), respectively. Externalizing symptoms were assessed in the four waves by mothers’ ratings of 10 items from the SDQ (Goodman, 1997). The 10 items were taken from the hyperactivity subscale (5 items) and the conduct problems subscale (5 items).

#### Socioeconomic status

SES was calculated from mother reports when children were 3 based on the number of rooms/number of residents’ ratio, income (reported on a scale from 1-considerably below average to 5-considerably above average, with 3 representing the national average), and mother’s years of education. These variables were standardized and then averaged (e.g., Avinun & Knafo-Noam, 2017).

#### Pregnancy and delivery information

When the twins were 3 and 5, mothers provided details regarding complications during pregnancy and/or delivery (e.g., rubella, RH mismatch, bleeding during pregnancy or miscarriage risk, preeclampsia, gestational diabetes, high risk pregnancy/bed rest, hospitalizations, umbilical cord prolapse, breech birth, assisted delivery, and placental abruption) and neonatal problems of each twin (neonatal hepatitis, neonatal diabetes, and being in an incubator). Complications during pregnancy/delivery and neonatal problems were summary scores of all relevant yes/no questions. Additional information that we included in our analyses: total weeks of pregnancy, Apgar scores of each twin 1- and 5-minutes after birth, whether each twin was hospitalized in the first year of life or afterwards, and breastfeeding (whether they breastfed each twin). Notably, maternal recall 4 to 6 years later regarding perinatal events has been found to be 89% accurate in comparison with hospital records (Githens, Glass, Sloan, & Entman, 1993).

#### Developmental problems

At all ages (i.e., 3, 5, 6.5, and 8-9) mothers were asked whether either twin suffered from developmental problems (e.g., stammering, speech delay, emotional, social, or communication problems, hypotonia or hypertonia, difficulty with gross/fine motor skills, behavior problems, hearing/vision impairment, attention/hyperactivity problems, eating problems, and wetting/toilet training problems). This was coded as 0 or 1.

#### Cognitive ability

The Block Design (visual processing) and the Vocabulary (crystallized intelligence) subsets of the Wechsler Intelligence Scale for Children-IV (WISC-IV; Lerner & Miller, 1978) were used to assess cognitive ability. The Block design subset includes 14 items and participants are asked to assemble blocks under a time limit, based on a pattern that is shown by the examiner. The Block design is scored based on accuracy and time. The vocabulary subset includes 37 items and participants are asked to explain a word using their own words. Scores reflect the accuracy of their definition. A mean of the two subsets’ scores was used as a cognitive ability score. Both subsets were administered at the lab or at the children’s homes when the twins were 6.5 years old.

#### Personality

At age 11 the twins were asked to fill in the Big Five Inventory (BFI) questionnaire (John, Donahue, & Kentle, 1991) either online or by hand. The BFI includes 44 items (e.g., “I am sometimes rude to others”) used to assess individual differences along the five-factor model of personality: neuroticism, extraversion, openness, conscientiousness, and agreeableness. Each scale was measured using 8-9 items, and an average requiring at least 5 non-missing items was calculated for each trait. Data was available for 1,349 children.

#### Self-Esteem

At age 11 the twins were asked to fill in the Rosenberg Self-Esteem Scale (Rosenberg, 1965) either online or by hand. The 10 items of the questionnaire (e.g., “I wish I could have more respect for myself”) were summed to create a self-esteem score. Data was available for 1,106 children.

#### Life satisfaction

At age 13 the twins were asked to fill in 4 items (”my life is going well”, “my life is just right”, “I have a good life”, and “ I have what I want in life”) taken from the Student’s Life Satisfaction Scale (Huebner, 1991). A mean of the 4 items (with a minimum of 3 items) was used as a life satisfaction score. Data was available for 703 children.

### Statistical analyses

#### Replication of the p-factor/Verifying the bifactor model

The same 20 items representing internalizing and externalizing symptoms were used for the factor analyses at each age, to allow developmental analyses and unambiguous comparisons across waves (as previously done, e.g., Murray, Eisner, & Ribeaud, 2016). The items were defined as categorical, which is needed in Mplus to indicate that they should be treated as ordinal variables in the model. As previously done (Avinun et al., 2020; Caspi et al., 2014; Romer et al., 2018), we used confirmatory factor analysis to fit three structural models: 1) A bifactor/hierarchical model, which consists of a *p*-factor that loads on all items, and two additional factors that are allowed to correlate, each loading separately on either internalizing or externalizing symptom items; 2) A one factor model, which consists of a factor that loads on all items; and 3) A correlated factor model, which consists of only two factors, each loading separately on either internalizing or externalizing symptom items. All confirmatory factor analyses were performed in Mplus version 7 (Muthén & Muthén, 2007) using the weighted least squares means and variance adjusted (WLSMV) algorithm, which is appropriate for ordinal data, and type=complex with the ‘cluster’ option, to account for the structure of the data (i.e., twins nested within families). This was done for each age (i.e., 3, 5, 6.5, and 8-9) separately.

We assessed how well each of the three models (one factor, correlated factors, and bifactor model) fit the data using the comparative fit index (CFI), the Tucker-Lewis index (TLI), and the root-mean square error of approximation (RMSEA). CFI and TLI closer to 1 and RMSEA closer to 0 indicate good fit (Hu & Bentler, 1999).

#### Externally validating the psychopathology factors

To test the external validity of the psychopathology factors, we examined the association between the three factor scores and previously identified correlates of psychological functioning: sex, personality traits and cognitive ability. All analyses were done in R version 4.0.3 (R Core Team, 2020). The package ‘geepack’ (Halekoh, Højsgaard, & Yan, 2006) was used to conduct linear regression analyses with generalized estimating equations (GEE) with an “exchangeable” correlation structure, due to the cluster structure of the data (i.e., families). The factors were entered as independent variables in separate analyses, and either personality traits (age 11) or cognitive ability (age 6.5) were entered as the dependent variables. In the analyses of sex, sex was entered as the independent variable. All models were tested separately.

#### Longitudinal twin analysis/Cholesky decomposition model

To test stability and change of the genetic and environmental influences on the psychopathology factors from age 3 to age 8-9 a longitudinal twin analysis*/*Cholesky decomposition model was conducted. Analyses were done in R version 4.0.3 (R Core Team, 2020) with the “openMx” (Neale et al., 2016) and “umx” (Bates, Neale, & Maes, 2016) packages and in the model-fitting program Mx version 1.70a (Neale, Boker, Xie, & Maes, 2003). Twin studies take advantage of the genetic difference between MZ twins, who share 100% of their genes, and DZ twins, who share on average 50% of their genes. The twin design assumes that if MZ twins are more similar than DZ twins then the individual differences in the examined phenotype are genetically influenced. These genetic influences can be represented by additive (A) and non-additive/dominant (D) genetic effects. Similarity beyond these genetic influences is attributed to the environment the twins share (shared or common environmental effects, C), and any differences between the twins are ascribed to the non-shared environment, which also includes measurement error (E). In a model that only includes reared-together twins, it is not possible to estimate D and C simultaneously. Therefore, when the presence of the D component is suggested (i.e., the MZ correlation is more than twice as large than the DZ correlation), an ADE model needs to be estimated instead of an ACE model. When DZ correlations are negative, a model that also includes a contrast effect has been suggested (Saudino, Cherny, & Plomin, 2000), as will be explained below.

In a longitudinal genetic model, a heritability component named A1 is created that estimates the variance explained by genetic influences on the first *p*-factor (age 3). The contribution of A1 to the *p*-factors at later ages (i.e., 5, 6.5, and 8-9) is also estimated, providing an indication of the stability of genetic influences across early development. Similarly, a heritability component named A2 is created that assesses the variance explained by genetic influences that are not accounted for by A1 and that affect the second (i.e., age 5 *p*-factor) and successive waves (i.e., ages 6.5 and 8-9). This is done for each age and for each of the model’s components (e.g., A, C, and E). Notably, when certain components are estimated to be very low, constrained models, in which these components are equated to zero, are fitted (e.g., an AE or a DE model). A best fitting model can be chosen based on the lowest Akaike information criterion (AIC).

#### The psychopathology factors as predictors and Predicting the psychopathology factors

Associations between the psychopathology factors and related measures were also performed in R in GEE models as explained above in the *Validating the p-factor* section (when developmental problems at age 8-9 were tested as the dependent variable, a logistic regression with GEE was performed). Here, sex (coded as 1=males, 2=female), age, and SES at age 3 were used as covariates.

## Results

Descriptive statistics are available in Supporting Table 2.

### Replication of the p-factor/Verifying the bifactor model

At all four ages, the structural model that fit the data best and showed adequate to good fit was the bifactor model; this was evidenced by the highest CFI and TLI scores and the lowest RMSEA value among the three models (Supporting Table 3). Factor scores from the bifactor model at each age were extracted and used in following analyses. The items and their standardized loadings on each factor are presented in Supporting Table 3. The strength of the loading on each item was relatively consistent between early and middle childhood, so that items with higher loadings on the *p*-factor at age 3 had higher loadings in all 4 waves (the correlations between the factor loadings at each age ranged between .97 and .98).

We examined longitudinal invariance of the *p*-factor by constraining the *p*-factor loadings to equal across ages and comparing this nested model to the unconstrained model. As in the original models, all means were fixed at 0 and the variances were fixed at 1, for model identification and standardization of the factors. To compare the models, we employed the DIFFTEST option in Mplus, which enables a chi-square difference test analysis when the WLSMV estimators is used. The difference test was significant (χ2 diff (60)=138.66, p<.0001), however, the RMSEA (.018), CFI (.914), and TLI (.909) in the unconstrained model were similar to the RMSEA (.017), CFI (.919), and TLI (.915) of the nested model. Therefore, and taking into consideration that chi-square tests are considered overly conservative in large sample sizes, it can be concluded that the *p*-factor is relatively stable during childhood. This is also supported by the correlations between the *p*, externalizing, and internalizing factor scores across ages (shown in Figure 1A. For purposes of the figure, we limited the sample to one random twin in each family to avoid dependency and allow a simple presentation). The correlations between the *p*-factor at different ages were consistently moderate to high (r=.45-.68). Correlations between the internalizing factors (r=.37-.58) and between the externalizing factors (r=.34-.67) were of similar magnitude.

**Figure 1.**
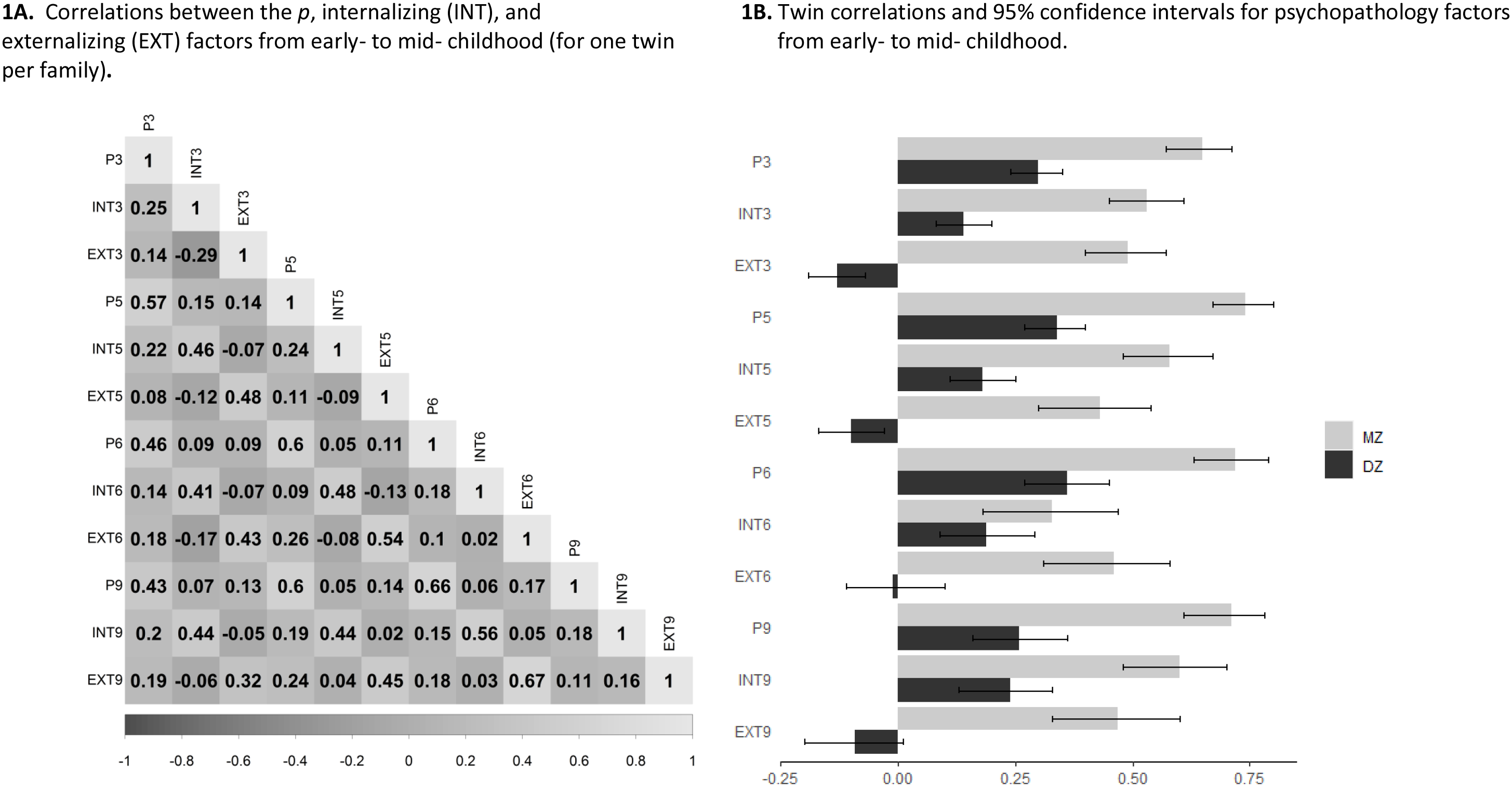
Psychopathology factors correlations.

### Externally validating the psychopathology factors

Patterns of association were examined across childhood between the psychopathology factors (*p*, externalizing, and internalizing), the big five personality traits (self-reports), cognitive ability (lab assessment), and sex. The psychopathology factors at each age were analyzed separately. Results are presented in Table 1 and Supporting Table 4. As expected, children with higher *p*-factor scores, tended to rate themselves as less agreeable, less conscientious, and more neurotic. Children with higher internalizing factor scores rated themselves as less extraverted and more neurotic, while children with higher externalizing factor scores rated themselves as more extraverted and less conscientious.

**Table 1.**
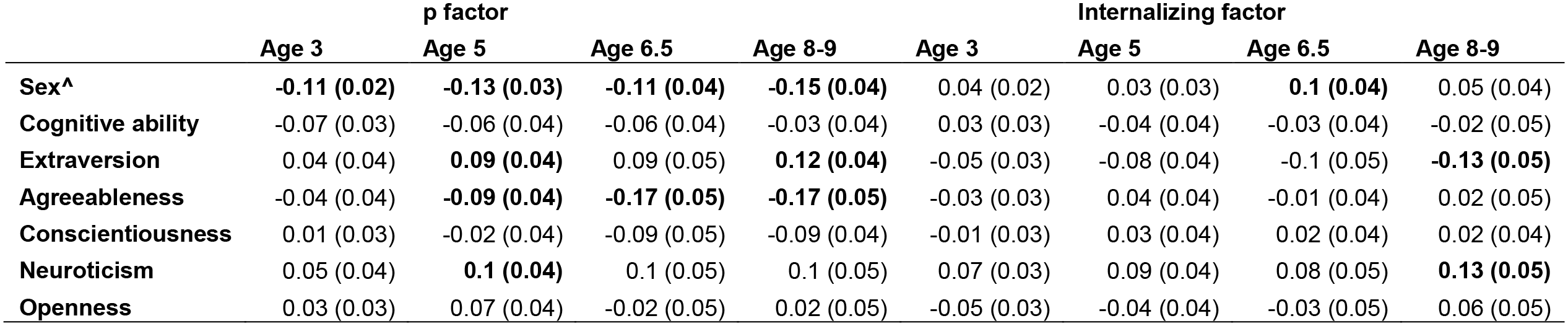
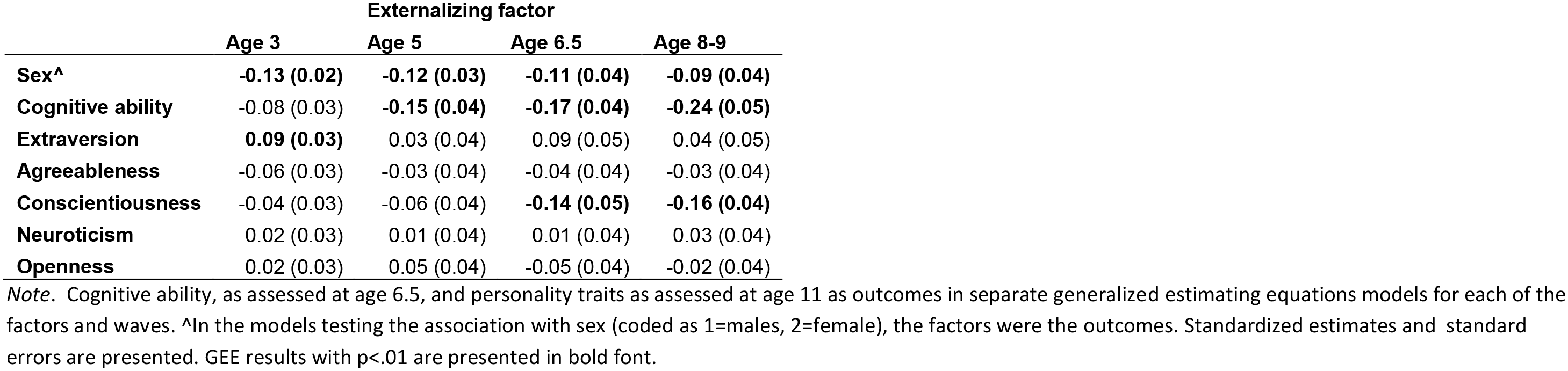
Associations between the three psychopathology factors, personality traits, cognitive ability, and sex from early to middle childhood. Standardized estimates and standard errors from generalized estimating equation models are presented.

Across ages 3 to 8-9, boys had higher *p*-factor scores than girls. Boys were also characterized by higher externalizing factor scores than girls, while girls tended to score higher on the internalizing factor, although this latter association was weak and inconsistent across ages. Cognitive ability, as assessed at age 6.5, showed the strongest negative association with the externalizing factor (R^2^ between .007 and .058), and only weak and inconsistent associations with *p* and the internalizing factor (R^2^ between 0 and .005).

### Longitudinal twin analysis/Cholesky decomposition model

Twin correlations for the psychopathology factors from early to middle childhood are presented in Figure 1B. MZ twin correlations were consistently higher than DZ twin correlations. Notably, in the case of the specific/residualized externalizing factor, DZ correlations were negative and weak, whereas MZ correlations were consistently moderate and ranged between .43 to .49. Sensitivity analyses showed that regressing the psychopathology factors on sex or excluding the DZ-opposite sex twins did not lead to meaningful changes in the correlations between MZ and DZ twins (Supporting Table 5).

For the *p*-factor the AE model fit the data best (Supporting Table 6) and is presented in Figure 2A (estimates with 95% confidence intervals are presented in Supporting Table 7A; because the difference in fit was small, an ADE model is also presented in Supporting Table 7B). The analysis indicated that the genetic influences on the *p*-factor are substantial and relatively stable in size (total heritability at each age ranged from .64 to .73), and that although new genetic influences arise at each age, a major proportion of the genetic influence is already present at age 3, and persists throughout childhood.

**Figure 2.**
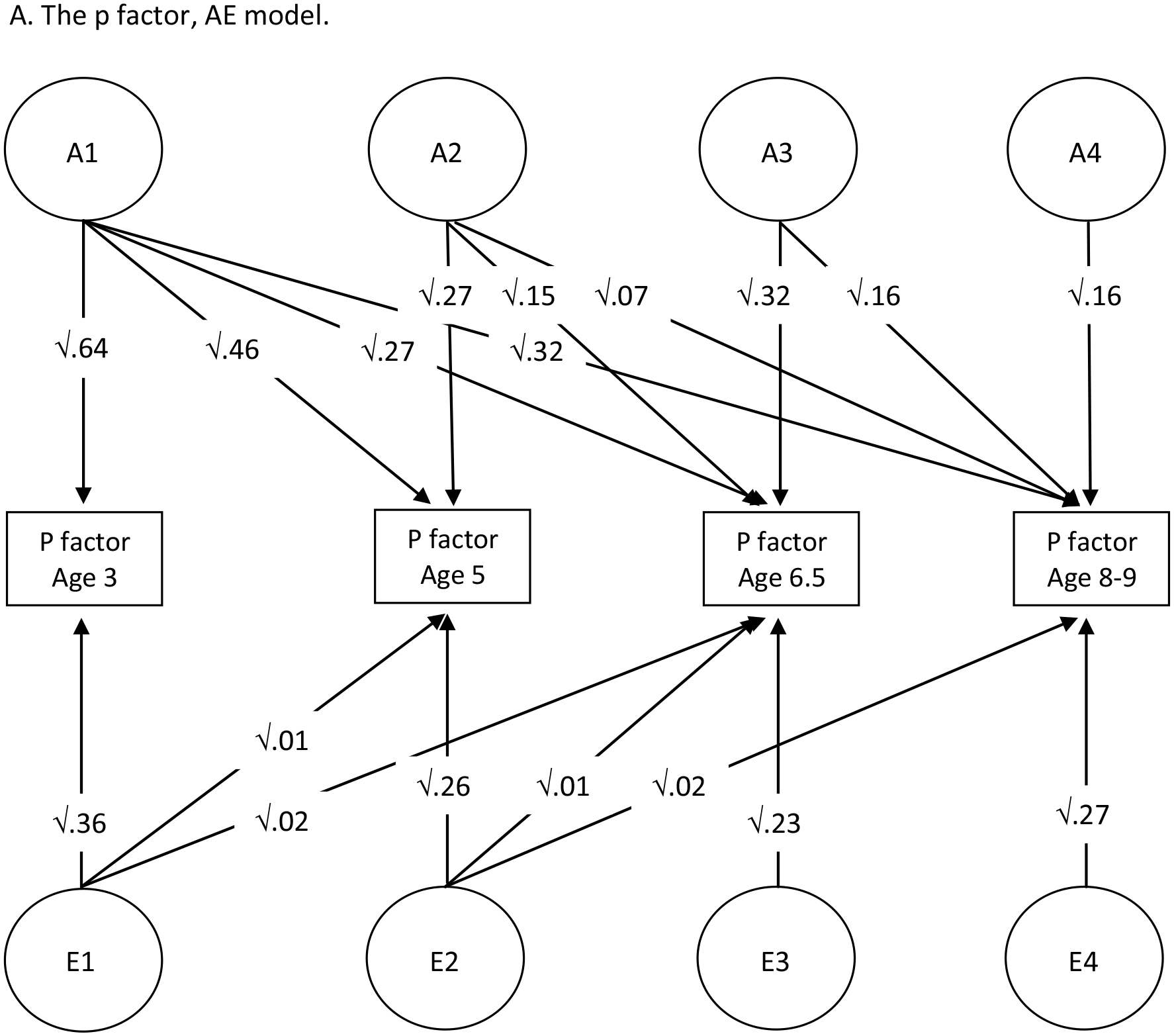
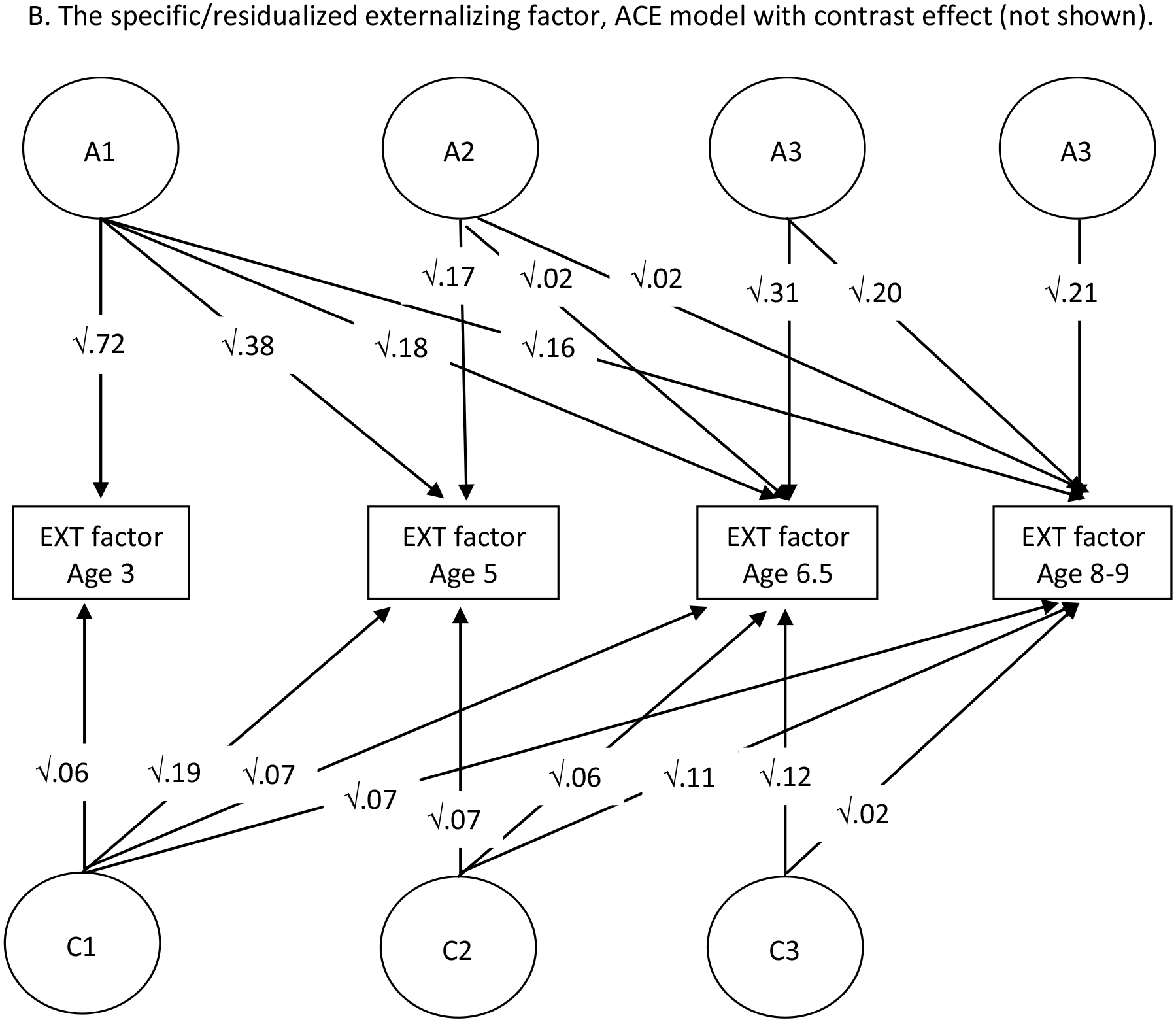
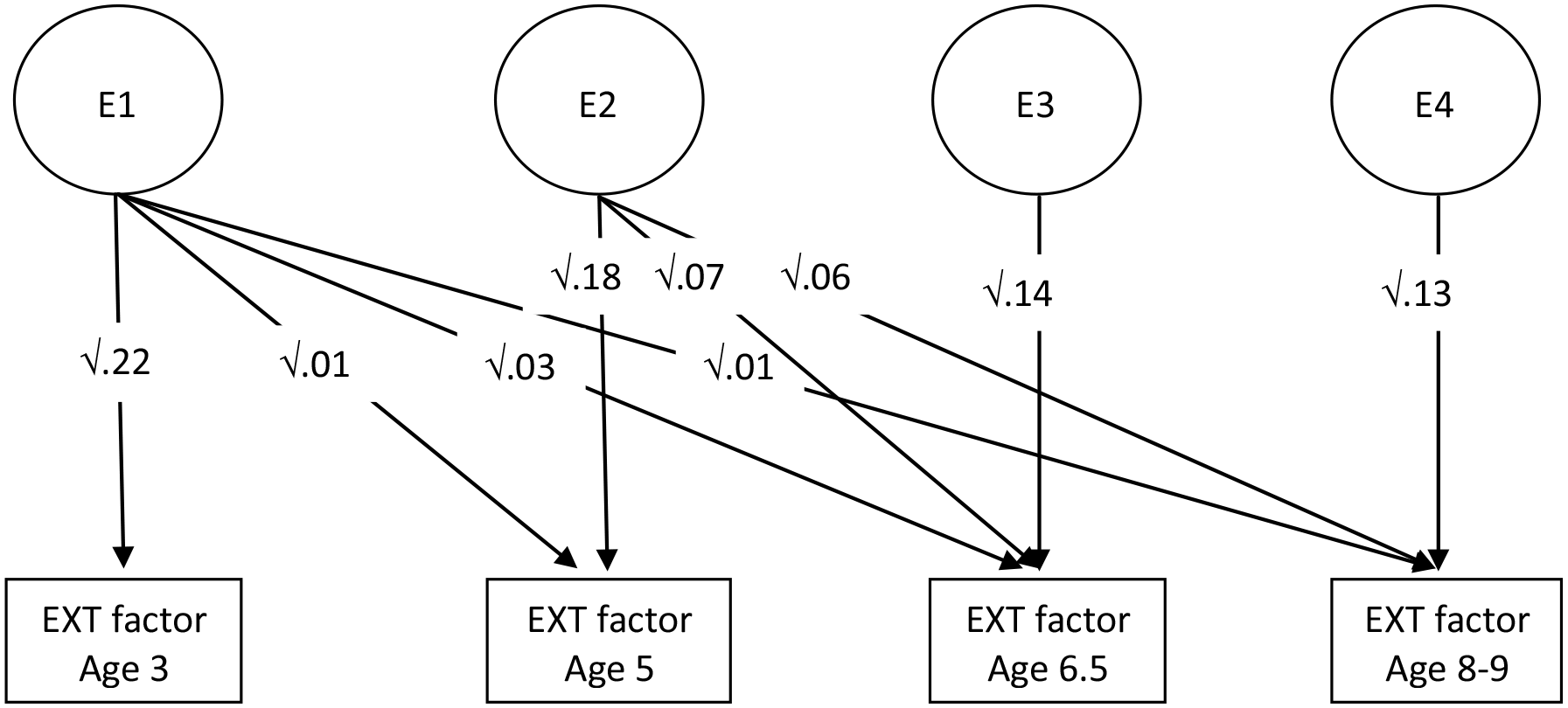
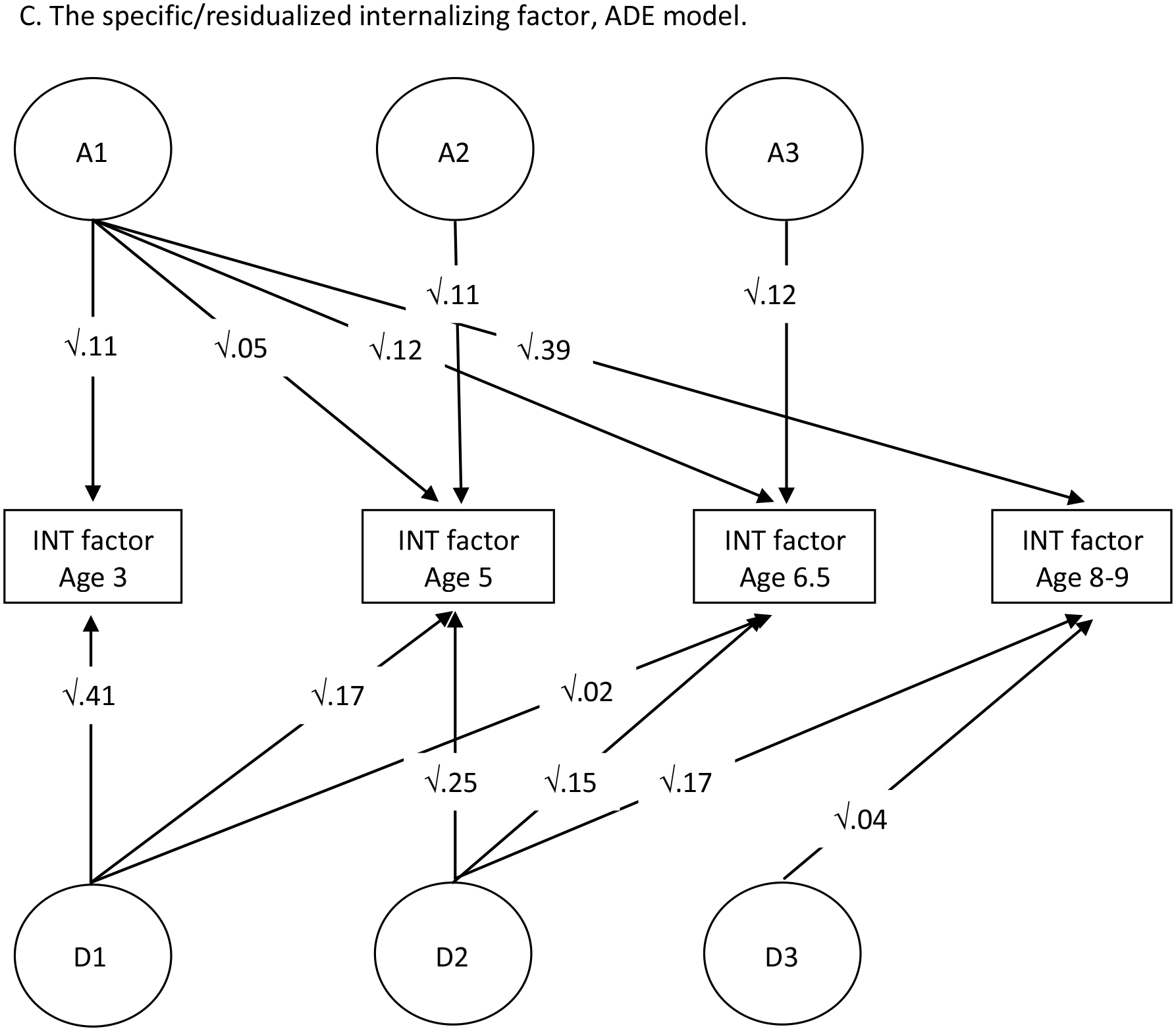
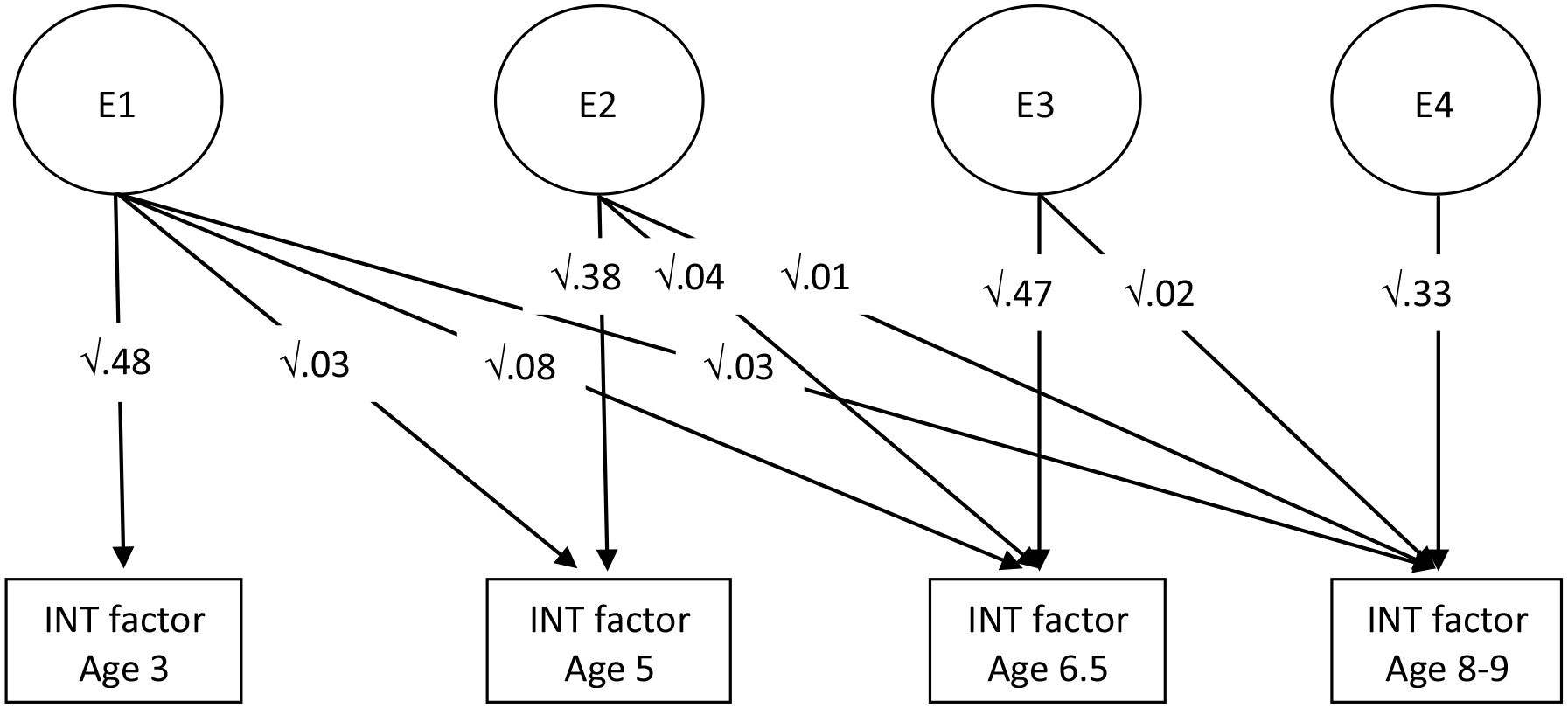
A longitudinal twin model. Genetic and environmental influences on psychopathology factors, from early to middle childhood.

As mentioned above, for the externalizing factor, DZ correlations were negative. Negative DZ twin correlations have been observed in previous studies (e.g., Goodman & Stevenson, 1989; Plomin et al., 1993), and specifically for hyperactivity (Thapar, Holmes, Poulton, & Harrington, 1999). Such correlations are thought to arise due to a contrast effect, which, in the case of negative DZ correlations, likely represents a bias in parent ratings (Saudino et al., 2000). In genetically influenced traits, DZ twins are expected to be more different than MZ twins. A contrast effect describes a situation in which parents exaggerate the existing difference between DZ twins. In addition to negative DZ correlations, a contrast effect is suggested when the DZs variance is greater than the MZs variance (Saudino et al., 2000). In our sample, the externalizing factor variance was consistently greater in DZs compared to MZs, although the differences were only significant in ages 3 and 5 (Supporting Table 8).

Based on the above, we opted to test a model for the externalizing factor that also accounts for a contrast effect (i.e., a path at each age from each twin’s phenotype to the phenotype of their co-twin. This path indicates that each twin’s phenotype is also a function of their co-twin’s phenotype) as shown in Saudino et al., (2000). Indeed, an ACE model with a contrast effect showed the best fit (Supporting Table 6). This model (without the contrast effect for simplicity of presentation) is shown in Figure 2B (estimates and contrast effects with 95% confidence intervals are presented in Supporting Table 9; the second best fitting model, the ADE model, is presented in Supporting Table 9B). The contrast effect at each age ranged between -.26 and -.32, suggesting the presence of parental bias. After accounting for this bias, the main portion of the variance across childhood was accounted for by genetic influences. The remaining variance tended to divide relatively evenly between the shared and non-shared environment. The genetic influences at age 3 continued to contribute to the variance across childhood. Notably, the shared environment confidence intervals were very large, suggesting the need to investigate these effects in a sample with greater statistical power.

For the internalizing factor, the DE model fit the data best (Supporting Table 6). However, a model with a dominant genetic effect (i.e., interaction effects), without an additive effect (i.e., main effect), is considered unlikely (Evans, 2020), and we therefore show the second best fitting model in Figure 2C, which is the ADE model (estimates with 95% confidence intervals are presented in Supporting Table 10A, and the DE model is presented in Supporting Table 10B). The ADE model for the internalizing factor suggested that about half (.41-.60) of the individual differences in internalizing symptoms from early to middle childhood are affected by genetic influences (whether additive or dominant). Most of these genetic effects were already present at age 3 or 5, suggesting that a large proportion of the genetic influences on internalizing symptoms is stable from early to middle childhood. The rest of the variance was accounted for by non-shared environmental effects and measurement error.

### The psychopathology factors as predictors

The *p*-factor was the most consistent and reliable of the three psychopathology factors in predicting developmental problems at age 8-9 (odds ratios ranging between 1.42 and 1.84; see Figure 3 and additional statistics in Supporting Table 11A). Interestingly, the *p*-factor retained its value as a predictor, as early as age 3, even when developmental problems from the same age as the *p*-factor were included in the model (odds ratios ranging between 1.36 and 1.60; Figure 3 and Supporting Table 11B).

**Figure 3.**
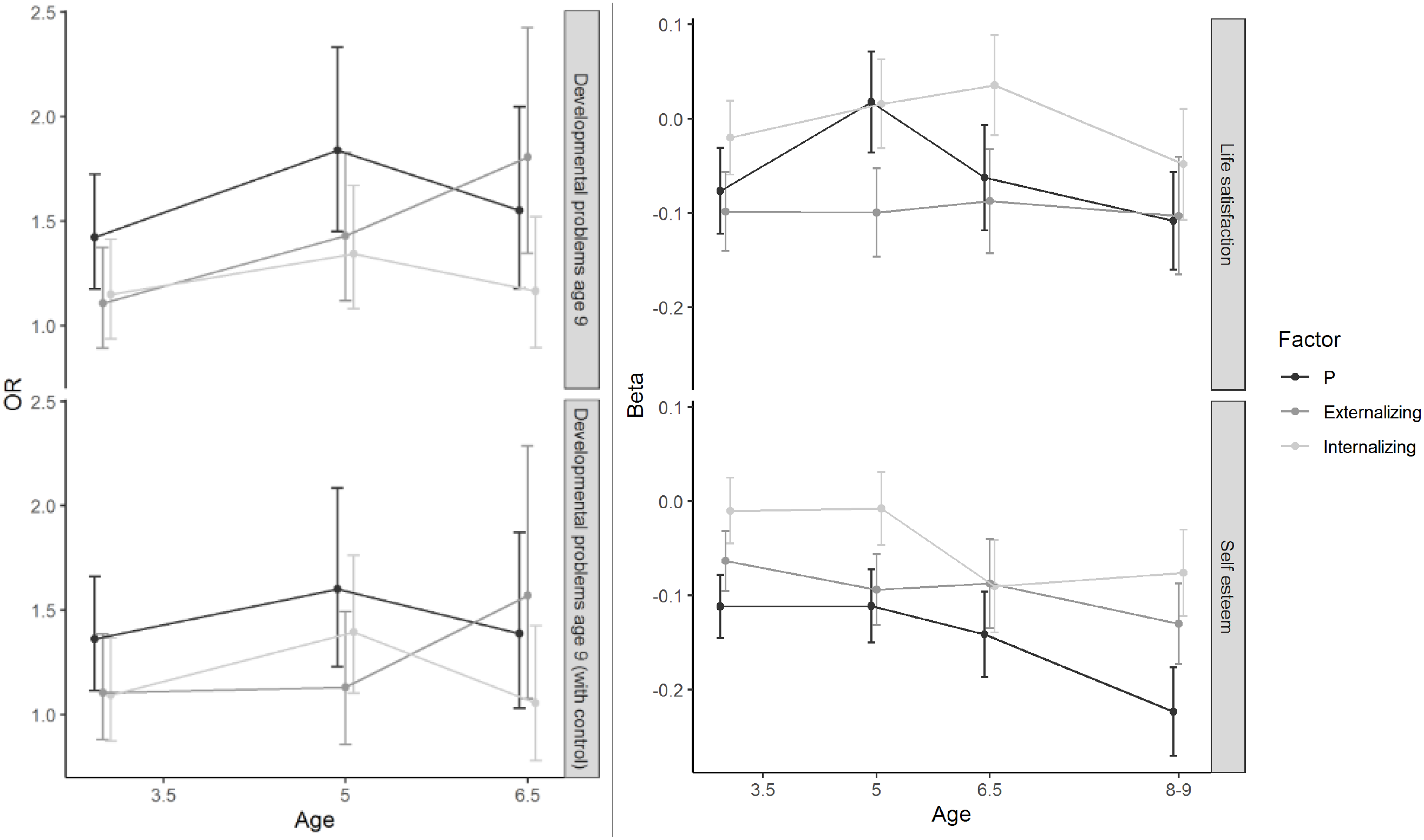
The psychopathology factors as predictors of developmental problems, self-esteem, and well-being, including error bars. *Note*. In all models age, sex (coded as 1=males, 2-female), and socioeconomic status as assessed at age 3 were entered as covariates. Results of generalized estimating equation models are presented. All models are independent (i.e., each factor was examined as a predictor in an independent statistical model). The model “with control” also included a developmental problems (yes/no) measure, assessed at the same age as the tested factor, as a covariate. Error bars are based on the standard errors.

The *p*-factor was also the best predictor of self-esteem at age 11 (R^2^ ranging from .015 to .04), indicating that, on average, children with higher *p*-factor scores reported lower self-esteem at age 11. Interestingly, the externalizing factor was the best predictor of life satisfaction at age 13 (R^2^ ranging from .007 to .014); children with higher externalizing symptoms tended to report poorer life satisfaction (Figure 3 and Supporting Table 11C).

### Predicting the psychopathology factors

Exploratory analyses, examining possible early-life predictors of the psychopathology factors, are presented in Table 2 and Supporting Table 12, which includes additional information such as p-values corrected for multiple comparisons. Most predictors explained about 1% or less of the variance in the psychopathology factors. On average across waves, the summary score of neonatal problems, which consisted of neonatal hepatitis, neonatal diabetes, and having been in an incubator, explained the most *p*-factor variance at each age (R^2^ from .004 to .013), such that greater neonatal problems were associated with higher *p*-factor scores across childhood.

**Table 2.**
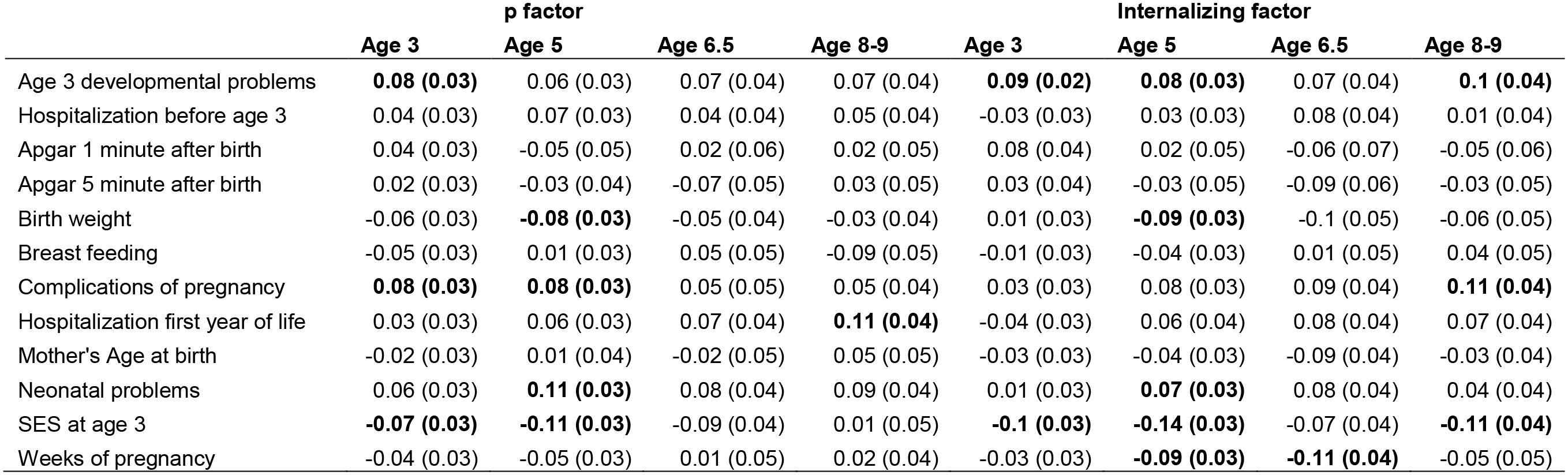
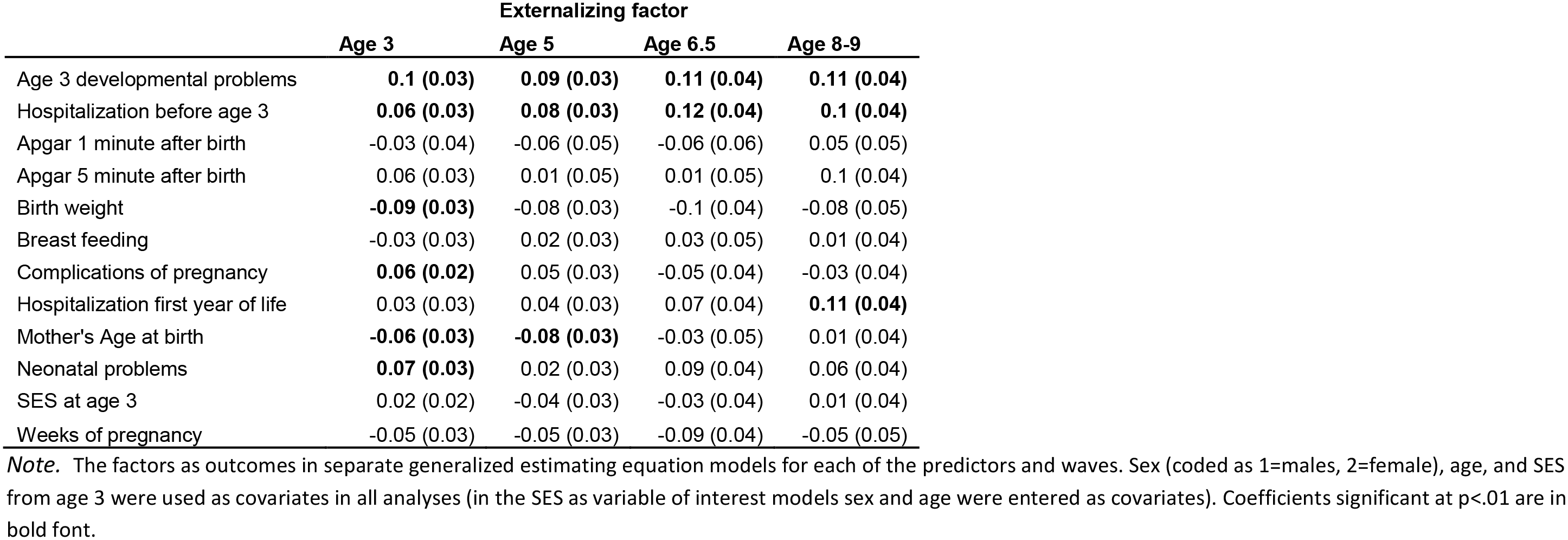
Possible predictors of the psychopathology factors from early to middle childhood.

|  | p factor |  |  |  | Internalizing factor |  |  |  |
| --- | --- | --- | --- | --- | --- | --- | --- | --- |
|  | Age 3 | Age 5 | Age 6.5 | Age 8-9 | Age 3 | Age 5 | Age 6.5 | Age 8-9 |
| Age 3 developmental problems | **0.08 (0.03)** | 0.06 (0.03) | 0.07 (0.04) | 0.07 (0.04) | **0.09 (0.02)** | **0.08 (0.03)** | 0.07 (0.04) | **0.1 (0.04)** |
| Hospitalization before age 3 | 0.04 (0.03) | 0.07 (0.03) | 0.04 (0.04) | 0.05 (0.04) | -0.03 (0.03) | 0.03 (0.03) | 0.08 (0.04) | 0.01 (0.04) |
| Apgar 1 minute after birth | 0.04 (0.03) | -0.05 (0.05) | 0.02 (0.06) | 0.02 (0.05) | 0.08 (0.04) | 0.02 (0.05) | -0.06 (0.07) | -0.05 (0.06) |
| Apgar 5 minute after birth | 0.02 (0.03) | -0.03 (0.04) | -0.07 (0.05) | 0.03 (0.05) | 0.03 (0.04) | -0.03 (0.05) | -0.09 (0.06) | -0.03 (0.05) |
| Birth weight | -0.06 (0.03) | **-0.08 (0.03)** | -0.05 (0.04) | -0.03 (0.04) | 0.01 (0.03) | **-0.09 (0.03)** | -0.1 (0.05) | -0.06 (0.05) |
| Breast feeding | -0.05 (0.03) | 0.01 (0.03) | 0.05 (0.05) | -0.09 (0.05) | -0.01 (0.03) | -0.04 (0.03) | 0.01 (0.05) | 0.04 (0.05) |
| Complications of pregnancy | **0.08 (0.03)** | **0.08 (0.03)** | 0.05 (0.05) | 0.05 (0.04) | 0.03 (0.03) | 0.08 (0.03) | 0.09 (0.04) | **0.11 (0.04)** |
| Hospitalization first year of life | 0.03 (0.03) | 0.06 (0.03) | 0.07 (0.04) | **0.11 (0.04)** | -0.04 (0.03) | 0.06 (0.04) | 0.08 (0.04) | 0.07 (0.04) |
| Mother's Age at birth | -0.02 (0.03) | 0.01 (0.04) | -0.02 (0.05) | 0.05 (0.05) | -0.03 (0.03) | -0.04 (0.03) | -0.09 (0.04) | -0.03 (0.04) |
| Neonatal problems | 0.06 (0.03) | **0.11 (0.03)** | 0.08 (0.04) | 0.09 (0.04) | 0.01 (0.03) | **0.07 (0.03)** | 0.08 (0.04) | 0.04 (0.04) |
| SES at age 3 | **-0.07 (0.03)** | **-0.11 (0.03)** | -0.09 (0.04) | 0.01 (0.05) | **-0.1 (0.03)** | **-0.14 (0.03)** | -0.07 (0.04) | **-0.11 (0.04)** |
| Weeks of pregnancy | -0.04 (0.03) | -0.05 (0.03) | 0.01 (0.05) | 0.02 (0.04) | -0.03 (0.03) | **-0.09 (0.03)** | **-0.11 (0.04)** | -0.05 (0.05) |

**Table 2.** Continued.
|  | Externalizing factor |  |  |  |
| --- | --- | --- | --- | --- |
|  | Age 3 | Age 5 | Age 6.5 | Age 8-9 |
| Age 3 developmental problems | **0.1 (0.03)** | **0.09 (0.03)** | **0.11 (0.04)** | **0.11 (0.04)** |
| Hospitalization before age 3 | **0.06 (0.03)** | **0.08 (0.03)** | **0.12 (0.04)** | **0.1 (0.04)** |
| Apgar 1 minute after birth | -0.03 (0.04) | -0.06 (0.05) | -0.06 (0.06) | 0.05 (0.05) |
| Apgar 5 minute after birth | 0.06 (0.03) | 0.01 (0.05) | 0.01 (0.05) | 0.1 (0.04) |
| Birth weight | **-0.09 (0.03)** | -0.08 (0.03) | -0.1 (0.04) | -0.08 (0.05) |
| Breast feeding | -0.03 (0.03) | 0.02 (0.03) | 0.03 (0.05) | 0.01 (0.04) |
| Complications of pregnancy | **0.06 (0.02)** | 0.05 (0.03) | -0.05 (0.04) | -0.03 (0.04) |
| Hospitalization first year of life | 0.03 (0.03) | 0.04 (0.03) | 0.07 (0.04) | **0.11 (0.04)** |
| Mother's Age at birth | **-0.06 (0.03)** | **-0.08 (0.03)** | -0.03 (0.05) | 0.01 (0.04) |
| Neonatal problems | **0.07 (0.03)** | 0.02 (0.03) | 0.09 (0.04) | 0.06 (0.04) |
| SES at age 3 | 0.02 (0.02) | -0.04 (0.03) | -0.03 (0.04) | 0.01 (0.04) |
| Weeks of pregnancy | -0.05 (0.03) | -0.05 (0.03) | -0.09 (0.04) | -0.05 (0.05) |

*Note.* The factors as outcomes in separate generalized estimating equation models for each of the predictors and waves. Sex (coded as 1=males, 2=female), age, and SES from age 3 were used as covariates in all analyses (in the SES as variable of interest models sex and age were entered as covariates). Coefficients significant at $p < .01$ are in bold font.

For the internalizing factor, family SES at age 3 appeared to be the strongest predictor, explaining the most variance on average across waves (R^2^ ranging from .005 to .017); children raised in families with higher SES tended to have fewer internalizing symptoms. For the externalizing factor, developmental problems at age 3 explained the most variance on average across waves (R^2^ ranging from .006 to .01); children with more developmental problems at age 3 tended to have more externalizing symptoms across childhood.

## Discussion

In the current study, we investigated the *p*-factor and the more specific/residualized internalizing and externalizing factors from early to middle childhood. In general, we found that 1) the *p*-factor is present in children as young as 3 years old; 2) there is little to no effect of the shared-environment for *p* and the specific/residualized internalizing factor, and all factors show strong and relatively stable genetic influences; 3) early life measure, such as childhood SES and neonatal and pregnancy risk, while at times significantly associated with the *p*-factor, do not explain more than 1% of the variance in any of the three psychopathology factors; and 4) the *p*-factor in early childhood can help to predict developmental problems and self-esteem in ages 8-9 and 11, respectively.

Our analyses indicated that a bifactor model which includes a general psychopathology factor, i.e., the *p*-factor, and two specific/residualized factors (internalizing and externalizing), fit the data best across all four waves from early to middle childhood. This supports and replicates previous findings of a transdiagnostic factor in childhood (e.g., McElroy, Belsky, Carragher, Fearon, & Patalay, 2018; Morales et al., 2021). However, the use of fit indices for determining the model that best represents the structure of psychopathology has been criticized, as simulations have shown that the bifactor model is found as the best fitting model even when simulated data is created based on a correlated factors model (Greene et al., 2019). We were therefore interested in externally validating the *p*-factor, by examining its association with known correlates.

Indeed, as previous studies on different samples have indicated (Avinun et al., 2020; Caspi et al., 2014; Etkin et al., 2020), the *p*-factor is negatively associated with agreeableness and conscientiousness, and positively associated with neuroticism. It is noteworthy that in our study the *p*-factor was estimated based on mother reports, while personality traits were assessed by self-reports of the children, thus ruling out shared method (source) variance as accounting for this association. The externalizing factor (net of *p*) was negatively associated with conscientiousness and positively associated with extraversion. In contrast, the internalizing factor (net of *p*) was positively associated with neuroticism and negatively associated with extraversion. This association between the three psychopathology factors and profiles of Big Five traits has now been demonstrated across cohorts varying in age and culture, and further supports the notion of overlapping taxonomies between the basic structure of personality and psychopathology (Brandes, Herzhoff, Smack, & Tackett, 2019).

We found that the externalizing factor (net of *p*) was most strongly associated with cognitive ability, a finding that is inconsistent with previous studies in which the *p*-factor was more strongly associated with measures of executive functioning (Caspi & Moffitt, 2018; Martel et al., 2017). It is possible that the use of different measurements or the reliance on symptom-level (our study) rather than diagnosis-level measures for the factor analysis (symptom counts or diagnostic probabilistic bands; Caspi & Moffitt, 2018; Martel et al., 2017) account for this inconsistency. Alternatively, if psychopathology is causal to cognitive impairment, it may be that disruptions to cognitive ability have yet to emerge in our young sample. Lastly, the items with the highest loadings on the externalizing factor related to inattention (e.g., easily distracted and cannot concentrate or finish tasks) which likely affected the children’s performance during the cognitive ability tasks.

The predictive utility of the *p*-factor was demonstrated by its positive association with developmental problems at age 8-9 and negative association with self-esteem, as measured in age 11 and self-reported by the children. While the specific developmental pathways leading to the latter association remain to be elucidated, one possibility may relate to previous findings showing that neuroticism, depression, and self-esteem are affected by shared genetic influences (Neiss, Stevenson, Legrand, Iacono, & Sedikides, 2009), suggesting that they have a shared etiology. Alternatively, psychopathology may increase the risk of exposure to stressful experiences, such as being bullied (Arseneault, Bowes, & Shakoor, 2010), leading to poorer self-esteem. Future research aimed at identifying these pathways has implications for how to most effectively target interventions.

Our finding of model invariance, as suggested by fit statistics, together with moderate to high correlations between the *p*-factor across waves, and relatively consistent factor loadings, which showed little change during development from early to middle childhood, indicated that *p* is reliable and stable across childhood. Additional support for the reliability of the *p*-factor was obtained from our longitudinal genetic analysis, which indicated that the *p*-factor was substantially heritable and that the genetic effects on the *p*-factor at age 3 meaningfully contribute to the genetic influences on the *p*-factor in all ages. The only previous longitudinal genetic study of the *p*-factor included twins from age 7 to 16 and has also found that the *p*-factor is highly heritable and that genetic factors contribute to stability (Allegrini et al., 2020).

Our longitudinal genetic analyses showed that genetic effects accounted for a large portion of the variance in the internalizing (.41-.60) and externalizing (.51-.72) factors. For the internalizing factor, both additive and dominant genetic influences (i.e., both main effects and interactions between genetic variants) were indicated, and there was no evidence for shared environment influences. The remaining variance was accounted for by the non-shared environment. For the externalizing factor a contrast effect was found, suggesting that parents tend to exaggerate the differences in externalizing symptoms between DZ twins. Thus, future studies should use observational methods or interviews to study the longitudinal genetic and environmental effects on the specific/residualized factors. Variance in the externalizing factor (net of *p*) not accounted for by genetic influences was evenly divided between the shared and non-shared environment. Importantly, the broad confidence intervals of the models for both specific/residualized factors, suggested that a larger sample with more statistical power may be needed.

Although the *p*-factor appears to represent a real shared general risk for psychopathology, its origins remain unclear. Here, we examined whether measures that can be assessed in early life, such as early childhood SES and pregnancy or neonatal complications, can predict the *p*-factor in childhood. This was guided by previous research suggesting that early childhood SES and fetal and early life programming may be linked to mental health (Lewis, Galbally, Gannon, & Symeonides, 2014; O’Donnell & Meaney, 2017; Peverill et al., 2020). All measures showed only weak associations, if at all, with the *p*-factor, explaining 1%, and usually less, of the factor variance. This was also true for the specific/residualized externalizing and internalizing factors. Similarly, developmental problems at age 3 did not explain more than 1% of the variance in the psychopathology factors. Taken together, these suggest that a cumulative risk factor score composed of many fetal and early life stressors may be a better approach for the prediction of the latent factors of psychopathology (Meehan et al., 2020).

### Strengths and Limitations

To our knowledge, the current study is the first to provide an in-depth developmental and longitudinal genetic investigation of the specific/residualized externalizing and internalizing factors from the bifactor model and the first to examine the links between the general and specific/residualized factors of psychopathology and pregnancy, obstetric, and neonatal measures. However, this study also has several limitations. First, the assessment of psychopathology was restricted to maternal reports and relied on a relatively limited number of items. While this is not exceptional (e.g., McElroy et al., 2018; Patalay et al., 2015), further research is needed to understand how these and the inclusion of only identical items across ages, affected the findings. Second, our sample consisted only of twins and therefore the generalizability of the associations between the psychopathology factors and other measures need to be replicated. Notably however, in the context of the associations with the pregnancy, obstetric, and neonatal measures, the reliance on twins was advantageous, as twin pregnancies are associated with higher complications compared to singletons (Obiechina, Okolie, Eleje, Okechukwu, & Anemeje, 2011), allowing for greater variance. Third, our study was restricted to the developmental period of early to mid-childhood; research encompassing longer developmental periods is needed.

### Conclusions

Our study supports accumulating research indicating a general factor of psychopathology, which represents transdiagnostic risk across mental disorders (Caspi & Moffitt, 2018), and shows that this general factor is highly heritable, discernible in early childhood, and stable from early to mid-childhood. Results also suggest that the *p*-factor is not meaningfully predicted by pregnancy, obstetric, and neonatal events. Lastly, to our knowledge, this is the first in-depth genetically informed investigation of the specific/residualized factors of psychopathology in childhood. Findings indicated moderate to large genetic influences on both specific/residualized factors with a meaningful portion of these influences already present at age 3.

## Data Availability

Data is available upon request due to ethical considerations.

## Acknowledgements

We thank the participating families for their cooperation and all the lab members of the Social Development lab throughout the years for data collection and coding. We would also like to thank Prof. Kimberly Saudino for her help with the contrast effect analyses. The Longitudinal Israeli Study of Twins (LIST) was founded by grant No. 31/06 from the Israel Science Foundation. The work was further supported by grant No. 1670/13 from the Israel Science Foundation, starting grant No. 240994 from the European Research Council, and by a grant from The Science of Generosity Initiative, funded by the John Templeton Foundation to Ariel Knafo-Noam. RA is supported by a Lady Davis fellowship. The authors declare they have no conflicts of interest.

## Notes

### Competing Interest Statement

The authors have declared no competing interest.

### Author Declarations

The protocol for the experiment was approved by the ethics committee of the Sarah Herzog Hospital, Jerusalem, and informed consent was obtained from all participating parents.

### Summary of Updates

The externalizing factor was reanalyzed in a twin model that includes a contrast effect and longitudinal invariance analyses were added

